# Enumerating asymptomatic COVID-19 cases and estimating SARS-CoV-2 fecal shedding rates via wastewater-based epidemiology

**DOI:** 10.1101/2021.04.16.21255638

**Authors:** Bradley W. Schmitz, Gabriel K. Innes, Sarah M. Prasek, Walter Q. Betancourt, Erika R. Stark, Aidan R. Foster, Alison G. Abraham, Charles P. Gerba, Ian L. Pepper

## Abstract

Wastewater-based epidemiology (WBE) was utilized to monitor SARS-CoV-2 RNA in sewage collected from manholes specific to individual student dormitories (dorms) at the University of Arizona in the fall semester of 2020, which led to successful identification and reduction of transmission events. Positive wastewater samples triggered clinical testing of almost all residents within that dorm; thus, SARS-CoV-2 infected individuals were identified regardless of symptom expression. This current study examined clinical testing data to determine the abundance of asymptomatic versus symptomatic cases in these defined communities. Nasal and nasopharyngeal swab samples processed via antigen and PCR tests indicated that 79.2% of SARS-CoV-2 infections were asymptomatic, and only 20.8% of positive cases reported COVID- 19 symptoms at the time of testing. Clinical data was paired with corresponding wastewater virus concentrations, which enabled calculation of viral shedding rates in feces per infected person(s). Mean shedding rates averaged from positive wastewater samples across all dorms were 6.84 ± 0.77 log10 genome copies per gram of feces (gc/g-feces) based on the N1 gene and 7.74 ± 0.53 log10 gc/g-feces based on the N2 gene. Quantification of SARS-CoV-2 fecal shedding rates from infected persons has been the critical missing component necessary for WBE models to measure and predict SARS-CoV-2 infection prevalence in communities. The findings from this study can be utilized to create models that can be used to inform public health prevention and response actions.

**Highlights:**

- Wastewater-based epidemiology with clinical testing monitored SARS-CoV-2 in dorms.
- 79.2% of SARS-CoV-2 infections were asymptomatic, and 20.8% were symptomatic.
- Clinical and wastewater data aggregated to estimate SARS-CoV-2 fecal shedding rate.
- Mean fecal shedding rate based on the N1 gene was 6.84 ± 0.77 log10 gc/g-feces.
- Mean fecal shedding rate based on the N2 gene was 7.74 ± 0.53 log10 gc/g-feces.

**Graphical Abstract:** 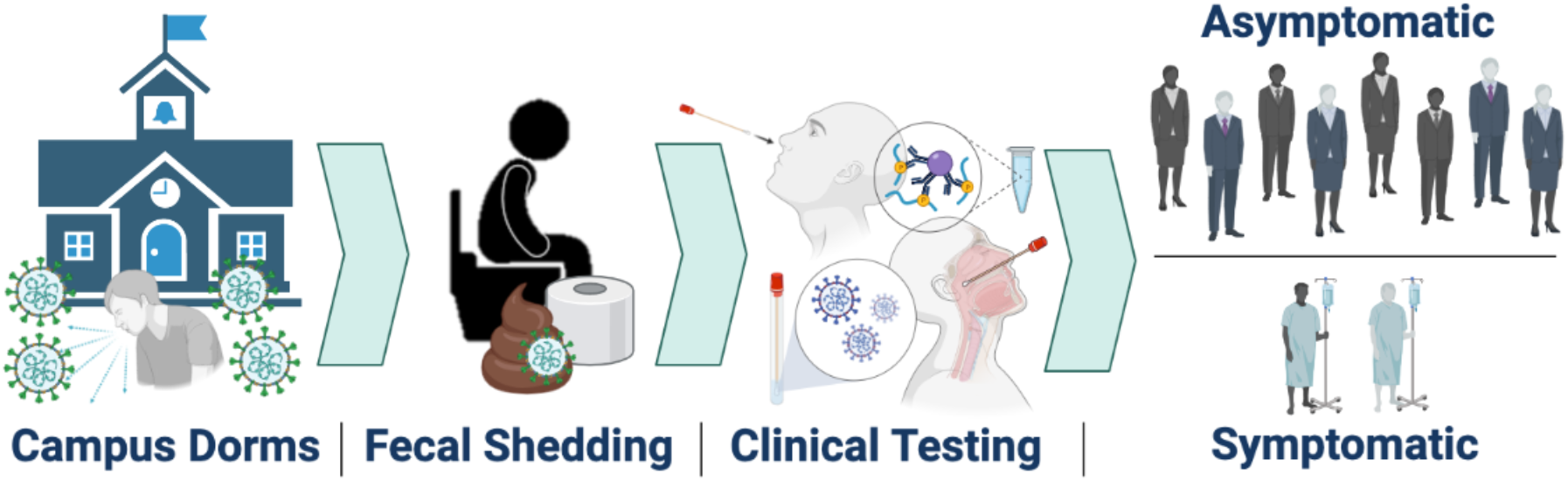

## 1. Introduction

SARS-CoV-2 infections may not end with current vaccination efforts. Some scientists conjecture that the virus’ evolution may require long-term monitoring and more intensive public health measures to control transmission of new variants. Thus, additional public health surveillance strategies can and should be implemented to prevent and respond to outbreaks; wastewater based epidemiology (WBE) may be one of those tools.

Human wastewater (i.e., sewage) may be among the first indications of aggregated, population-based SARS-CoV-2 infections in a community (Medema et al., 2020b), followed by healthcare facility reported number of infections, hospitalizations, and ultimately deaths. WBE is an effective, critical tool that has been demonstrated to monitor concentrations of SARS-CoV-2 RNA in wastewater as an indicator to survey the SARS-CoV-2 infection prevalence at a population level (Medema et al., 2020b). The results from these surveys can be used to track community infection dynamics (Peccia et al., 2020) and guide targeted public health response actions (Betancourt et al., 2021). Therefore, fecal shedding of SARS-CoV-2 may be utilized as a biomarker to estimate SARS-CoV-2 infections in a population..

Shedding estimates of SARS-CoV-2 from infected individuals have strong implications for the effectiveness of WBE to determine disease prevalence and guide public health interventions. Although SARS-CoV-2 has been shown to shed in the upper respiratory tract, the lower respiratory tract, feces, urine and serum (Cevik et al., 2020), WBE leverages RNA shedding of SARS-CoV-2 in feces. Infected persons shed virus into sewage via feces (Cevik et al., 2020), or urine (Brönimann et al., 2020), both of which can then be used to monitor population-level infection changes. Although SARS-CoV-2 has been isolated in urine of a COVID-19 patient (Sun et al., 2020), the incidence of the virus in urine is reported to be low (Brönimann et al., 2020; Morone et al., 2020); whereas, intact SARS-CoV-2 virus have been isolated from feces (Wang et al., 2020; Xiao et al., 2020; Zhang et al., n.d.). Further, viral RNA has been recovered even in the absence of intact virus isolation from stool samples (Wölfel et al., 2020), highlighting the sensitivity of molecular methods applied for WBE.

During the early stages of the pandemic in the Netherlands, SARS-CoV-2 was detected in sewage six days before initial cases were reported (Medema et al., 2020a). Mulitple jurisdictions around the world have monitored concentrations of SARS-CoV-2 RNA in sewage samples to quantify the total number of infected persons in the community that excreted the virus into wastewater (Ahmed et al., 2020; Chavarria-Miró et al., 2021; Curtis et al., 2020). More recently, the University of Arizona (UArizona) used WBE, dovetailed with targeted clinical testing, to prevent COVID-19 outbreaks in student dormitories (dorms). In particular, 91 wastewater samples containing SARS-CoV-2 RNA provided early-warning that at least one infected individual was present in the community (Betancourt et al., 2021). The UArizona case study highlighted the effectiveness of WBE to detect the presence of SARS-CoV-2 infected individuals in a defined community and to ultimately contain outbreaks by triggering public health response actions. Although WBE has been utilized to raise the alert of SARS-CoV-2 presence with a dichotomous threshold, quantifying the number of SARS-CoV-2 infected individuals cannot be done without accurate fecal shedding rate estimates.

This research aims to estimate SARS-CoV-2 fecal shedding rates in infected persons by aggregating data from positive wastewater samples with numbers of known symptomatic and asymptomatic individuals in defined communities. To that end, the Campus Re-entry study at the UArizona in Fall 2020 (Betancourt et al., 2021) provided a unique setting to calculate mean shedding rates for defined communities. Specifically, 13 dorms served as a case study to test the utility of WBE to monitor SARS-CoV-2 levels in sewage and initiate public health action based upon those results. Positive SARS-CoV-2 detection in wastewater samples from any given dorm led to point prevalence antigen and/or PCR testing for the vast majority of dormitory residents regardless of symptomatic expression; thus, the numbers of symptomatic and asymptomatic individuals was known. This current study determines the proportion of COVID-19 infections that were asymptomatic versus symptomatic and pairs these clinical data with wastewater results to extrapolate the fecal shedding rate of SARS-CoV-2 in infected students. Findings from this study will inform how WBE can be utilized for public health prevention and response actions.

## 2. Methods

### 2.1. Dormitory Sites

In total, 13 dorms (Dorm A–M) were monitored throughout the Fall 2020 Semester (August 17–November 20). Data from the final weeks of classes and exams following the Thanksgiving break (November 21–December 17) are not included in this analysis, as students were advised to complete the semester virtually and not return to campus. Each wastewater sample was collected from sewer manholes specific to each dorm’s effluent prior to convergence with other sewer pipelines; thus, all wastewater samples were specific to the defined communities living in each dorm. However, two sets of dorms (Dorm E and F; Dorm H and I) required sampling from a single sewer manhole distal from the mixing of wastewater from the two dorms but proximal to convergence with other pipelines. The dorms in each set are located directly side-by-side and were considered a ‘combined dorm’ for all public health response actions (i.e., clinical testing interventions) and data analysis. Dorms varied in infrastructure and resident occupancy (Table 1). Due to the pandemic, UArizona limited dorm room occupancy to a maximum of two.

**Table 1:** Meta-data for student dormitories.

| **Fall 2020** |  |  |  |  |
| --- | --- | --- | --- | --- |
| **Dorm** | **Occupancy<sup>a</sup>** | **Capacity** | **Room Type** | **Bathrooms** |
| A | 611 | 722 | Single, Double | Community, All-Gender |
| B | 342 | 400 | Single, Double | Community |
| C | 123 | 181 | Single, Double, Triple<sup>b</sup> | Community |
|  |  |  | Single, Double, Suite- |  |
| D | 623 | 731 | Style | Suite |
| E | 206 | 300 | Single, Double | Community, All-Gender |
| F | 56 | 60 | Single | Community |
| G | 231 | 300 | Apartments | In-Room (1 per bedroom) |
| H | 195 | 238 | Single, Double | Community, All-Gender |
| I | 181 | 238 | Single, Double | Community, All-Gender |
| J | 424 | 482 | Single, Double | Community |
| K | 328 | 369 | Double | Community, All-Gender |
| L | 76 | 106 | Single, Double, Triple<sup>b</sup> | Community, Sink-in-Room |
| M | 132 | 152 | Single, Double | Community |

<sup>a</sup> Occupancy varied throughout the Fall 2020 semester, so the highest numbers of residents at any given time throughout the semester are shown.
<sup>b</sup> Due to the pandemic, UArizona limited rooms to a maximum of two occupants. No triple-occupant rooms were offered during the time of this analysis.

### 2.2. Wastewater Sampling and Analysis

Wastewater samples from each dorm were collected between 9:00 and 10:30 am, then analyzed for SARS-CoV-2 RNA at least twice per week. One set of dorms was monitored on Monday/Wednesday/Friday (Dorms A, B, C, D, G, L), while a second set was monitored Tuesday/Thursday/Saturday (Dorms E-F, H-I, J, K, M). Grab samples were collected inside sewer manholes with sterile Nalgene® bottles that were affixed to extension poles, which allowed collection at depths of 5 to 15 feet. Velocity (ft^3^/s) of wastewater within the sewer was measured using a Global Water FP 211 flow probe (Global Water, College Station, TX). Multiple readings of the minimum and maximum flow rates (gallons per minute) over a one-minute period were averaged and recorded. Flow rates were calculated using Manning’s equation for partially full pipes (Akgiray, 2005). For all wastewater samples, the wastewater’s height and depth was estimated to be approximately two inches, based upon the height of the fluid on the propellor sensor. Note, this assumption was necessary since exact measurements were not possible inside sewer manholes. The depth of the wastewater was never above the propellor sensor and a velocity reading could not be recorded if wastewater height was not near the top of the propellor (2-inch height) but was nonetheless considered reasonable. Statistical methods were used to estimate flow rates for samples for which velocity could not be measured due to site obstruction or the flow probe not being available (See Section 2.6).

Grab samples collected at the same time of each sampling day were considered adequate based on a prior multiple sampling event. In that case, multiple samples were collected at a particular site over a 30-minute period, and analysis indicated virtually identical virus concentrations (Betancourt et al., 2021). This suggest that virus particles disperse upon entering the piping/sewer and remain in the sewer for extended periods of time rather than being removed by plug flow (Manor et al., 2014). Samples were stored on ice and travel time to WEST was 30 minutes or less from the dorms. Wastewater processing and analysis for SARS-CoV-2 RNA followed procedures previously described (Betancourt et al., 2021). Samples were tested for the virus using the United States (U.S.) Centers for Disease Control and Prevention (CDC) RT-PCR assays that target regions of the nCoV nucleocapsid gene (N1 and N2; Table S1) (Research use only kit, Integrated DNA Technologies, Coralville, IA). Real-time PCR for the simultaneous detection of N1 and N2 genes was performed on a LightCycler 480 Instrument II (Roche Diagnostics) with the LightCycler® Multiplex RNA Virus Master (Roche Diagnostics) (Table S2).

### 2.3. Clinical Testing Data

Positive detection of SARS-CoV-2 RNA (via N1 and N2 gene regions) in wastewater was utilized as a leading indicator of the presence of SARS-CoV-2 infections within the dorm communities. Results were immediately communicated to the UA Task Force and Campus Re-Entry Working Groups, which planned and conducted clinical testing of residents as a response action(s), as previously described (Betancourt et al., 2021).

Clinical tests for COVID-19 diagnosis were performed by antigen testing via nasal swab samples and RT-PCR via nasopharyngeal swab samples (Betancourt et al., 2021). The analytical performance characteristics for the antigen test were 96% sensitivity and 100% specificity, as provided by the manufacturer (Sofia SARS Antigen FIA, Quidel, San Diego, CA, USA). Performance of the RT-PCR tests were determined by the University of Arizona Genetics Core for Clinical Services (CDC 2019-nCoV RT-PCR Diagnostic Panel) with a limit of detection at 150 viral copies/reaction, or 30 viral copies/μl of sample.

Clinical testing data was obtained from the University Campus Health Services (CHS) and the Test All Test Smart (TATS) program. Symptomatic and asymptomatic cases in the study were tracked based on location and program where clinical tests were conducted. Individuals with COVID-19 symptoms who sought clinical testing were tested through CHS. Clinical testing of non-self-reporting (i.e., asymptomatic and sub-clinical) students and employees was conducted through the TATS program that runs several testing locations on campus and mobilizes pop-up sites as needed (i.e., targeted clinical testing at dorms with positive wastewater detection of SARS-CoV-2). A negative antigen test at CHS, where patients were exhibiting symptoms, was followed by PCR testing to confirm results; data was de-duplicated in situations when both testing methods were used to confirm test results (Betancourt et al., 2021). To ensure compliance with the Human Subjects Protection Program (HSPP), the use of clinical data was reviewed and approved by a UArizona Institutional Review Board (IRB).

### 2.4. Alignment of wastewater and clinical data

To estimate the number of SARS-CoV-2 infected individuals who contributed to a single positive wastewater sample, a 6-day range of clinical data was considered. Positive clinical cases from the day before, day-of, and four days after sampling were included in the count of infected individuals contributing to viral shedding. The rationale for this approach is:

1) Residual virus—shed from individuals testing positive for COVID-19 the day prior to sampling—may be detected in wastewater the following day. Previous sampling indicates that virus can persist for extended periods of time and be detected even when an estimated 1000 gallons of wastewater has continued to flow through a sewer system (Betancourt et al., 2021).

2) Individuals infected with SARS-CoV-2 may shed virus into wastewater prior to showing symptoms and/or being identified as a clinical case. The viral load of SARS-CoV-2 appears to peak in infected persons within the first week of infection (Cevik et al., 2020) and the median incubation period for COVID-19 is estimated to be approximately five days (Lauer et al., 2020).

Following a positive wastewater sample, clinical testing was performed on nearly all residents living in the dorm via the TATS program (Betancourt et al., 2021). Individuals that tested positive were removed from the dorm and transferred into isolation. However, some individuals in the early stages of infection may have tested negative on the day of wastewater testing, but tested positive after reaching peak viral load of SARS-CoV-2 a few days later. It is possible that these individuals were shedding virus in feces even though they tested negative via initial TATS conducted within 24 hours of a positive wastewater signal. Additionally, the full dorm community was not always available on the first day of targeted testing and were tested a day or more after a positive wastewater detection. Therefore, clinical data for 4-days following positive wastewater samples was included in the count of infected individuals since these persons likely shed and contributed virus to the wastewater samples for several days.

The majority (68 out of 81) of positive wastewater samples were associated with new reported cases of infections within this 6-day period.

### 2.5. Viral Shedding Rate Estimation

The fecal shedding rate of SARS-CoV-2 RNA per gram of feces from an infected individual was enumerated based on known concentrations of viral RNA in positive wastewater samples and the fact that the total numbers of infected persons contributing to the total virus load in the samples were also known. Equations from previous reports using viral loads in wastewater to estimate the number of infected persons in a community (Ahmed et al., 2020; Chavarria-Miró et al., 2021; Curtis et al., 2020) were modified so that the fecal shedding rate of viral RNA per infected person could be quantified. The fecal shedding rate (FS) in terms of genome copies per gram-feces (gc/g-feces) was calculated as follows:

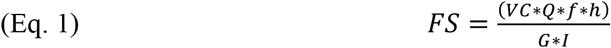

where VC is the virus concentration (genome copies/L) in the wastewater sample, *Q* is the flow rate (gpm) of wastewater in the sewer manhole at time of sample collection, *f* is the conversion factor between gallons and liters, *h* is the conversion factor between minutes and days, *G* is the typical mass of stool produced per person per day (Curtis et al., 2020; Rose et al., 2015), and *I* is total number of infected persons contributing to the wastewater sample based on the 6-day range for clinical data (see Section 2.5). Confidence intervals were calculated based on the standard deviations between sample calculations.

Fecal shedding rates based on N1 and N2 genes were calculated independently.

### 2.6. Statistical Analysis

Statistical analyses were performed in Microsoft Excel (version 16.47.1, 2021) and R studio (R Studio Team (2020). RStudio: Integrated Development for R. RStudio, PBC, Boston, MA. *http://www.rstudio.com/*). Multiple imputation for measurement error (MIME) was performed to estimate flow rates for missing data when the flow probe was unavailable, as previously described with modifications (Canales et al., 2018).

## 3. Results

### 3.1. Asymptomatic and Symptomatic Clinical COVID-19 Cases

Asymptomatic and symptomatic cases of COVID-19 were differentiated based on clinical testing program – TATS for asymptomatic and CHS for symptomatic (see Section 2.4). For all dates in the study period, a total of 711 clinical cases were reported among the 13 unique dorms. Clinical data was combined for dorms with converged sewage systems (Dorm E-F and Dorm H-I). Across all dorms, 148 symptomatic cases and 563 asymptomatic cases were reported; thus, 79.2% of SARS-CoV-2 infections were asymptomatic and only 20.8% were symptomatic (Table 2).

**Table 2:** Clinical positives for COVID-19 in wastewater-monitored dorms, 8/17/20-11/20/20

| **Dorm** | **Clinical**<br>**Positives** | **Symptomatic (CHS)** |  | **Asymptomatic (TATS)<sup>a</sup>** |  |
| --- | --- | --- | --- | --- | --- |
|  |  | **Total** | **Percent<sup>b</sup>** | **Total** | **Percent<sup>b</sup>** |
| A | 164 | 35 | 21.3% | 129 | 78.7% |
| B | 92 | 16 | 17.4% | 76 | 82.6% |
| C | 10 | 3 | 30.0% | 7 | 70.0% |
| D | 171 | 34 | 19.9% | 137 | 80.1% |
| E & F | 16 | 8 | 50.0% | 8 | 50.0% |
| G | 2 | 0 | 0.0% | 2 | 100.0% |
| H & I | 74 | 17 | 23.0% | 57 | 77.0% |
| J | 111 | 16 | 14.4% | 95 | 85.6% |
| K | 66 | 15 | 22.7% | 51 | 77.3% |
| L | 0 | 0 | - | 0 | - |
| M | 5 | 4 | 80.0% | 1 | 20.0% |
| Total | 711 | 148 | 20.8% | 563 | 79.2% |

<sup>a</sup> Asymptomatic cases may also include mildly symptomatic individuals that did not self-report.
<sup>b</sup> Percent of total new clinical positive cases of infection.

Examining specific dorm communities, reported clinical cases ranged from zero in Dorm L to 171 in Dorm D (Table 2). Dorms G and M had the fewest reported infections, as well as the highest and lowest asymptomatic rates. With limited data (two infections in Dorm G and five infections in Dorm M), the asymptomatic rate was calculated as 100% for Dorm G and 20% for Dorm M. Combined Dorm E & F was the only dorm to report equal number of symptomatic and asymptomatic cases (Table 2). Seven of the eleven monitored communities had asymptomatic rates between 70 – 85% (Table 2). Two dorms (E & F and M) had asymptomatic rates at 50% or below, while Dorm G was the only to report 100% of new cases as asymptomatic (Table 2).

### 3.2. Virus Fecal Shedding Estimation

In total, 81 samples in the study period were positive for SARS-CoV-2 N1 and/or N2 gene(s) and 238 samples resulted in no detection of the virus (Table S3). Clinical and wastewater data were aggregated (see Section 2.5) to calculate the N1 and N2 viral shedding rate per infected person (see Section 2.6). Only 13 positive wastewater samples were omitted from viral shedding calculations due to zero reported cases of infection within the 6-day range for clinical data (Table S3). Negative wastewater samples were not included in viral shedding estimations due to no detection of SARS-CoV-2 N1/N2 genes. Also, negative samples did not trigger a response action to conduct clinical testing on residents; thus, there is limited clinical data for the days that wastewater was negative. Overall, 68 total positive wastewater samples were aligned with clinical data to extrapolate the fecal shedding rates of SARS-CoV-2 N1 and/or N2 genes.

The average N1 shedding rate per infected person was calculated to be 6.84 ± 0.77 log10 gc/g-feces, considering all positive wastewater samples (n=60) across all dorms (Table 3). The median was 6.65 log10 gc/g-feces with a full range 5.74 - 9.76 log10 gc/g-feces (Table 3). Within specific defined communities, Dorm C had the highest average N1 shedding rate at 7.26 ± 0.14 (n=2), while Dorm A had the lowest average at 6.45 ± 0.60 log10 gc/g-feces (n=13). The widest range of N1 shedding rates was found in Dorm J with a minimum at 5.98 and a maximum at 9.76 log10 gc/g-feces.

**Table 3:**
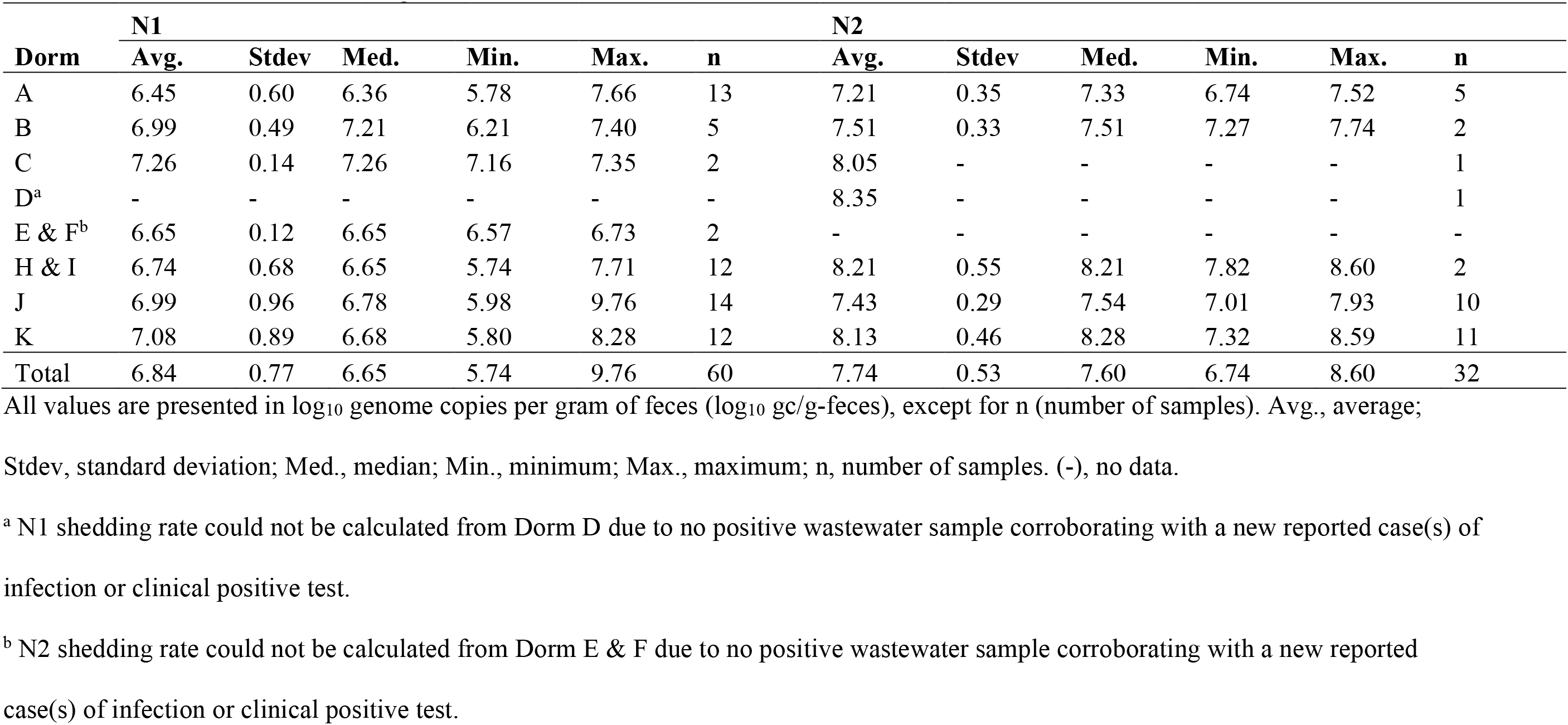
SARS-CoV-2 Viral Shedding Per Infected Person Extrapolated from UArizona Dorm WBE

The average N2 shedding rate per infected person was calculated to be 7.74 ± 0.53 log10 gc/g-feces across all positive wastewater samples (n=32) from all dorms (Table 3). The median was 7.60 with a full range from 6.74 – 8.60 log10 gc/g-feces (Table 3). For specific communities, combined Dorm H & I had the highest average N2 shedding rate at 8.21 ± 0.55 (n=2), while Dorm A had the lowest average at 7.21 ± 0.35 (n=5) log10 gc/g-feces. The widest range of N2 shedding rates was found in Dorm K with a minimum at 7.32 and a maximum at 8.28 log10 gc/g-feces.

With just one positive wastewater sample (n=1) each, Dorms C and D had the lowest number of estimates for N2 viral shedding rates (Table 3). For Dorm D, the N1 shedding rate could not be calculated due to zero wastewater samples being positive (i.e., non-detect) for the N1 gene target (Table S3D). Although Dorm D (8.35 log10 gc/g-feces) reported a higher N2 shedding rate than Dorm H & I (8.21 log10 gc/g-feces), it was not reported as a higher average due to only a single sample (n=1). Combined Dorm E & F also had a low number of samples with positive detection for N1 (n = 2) and none for N2 (Table 3; Table S3E); thus, the shedding rate for this combined dorm community could not be calculated for N2.

## 4. Discussion

### 4.1 Estimation of Asymptomatic Cases as a Percent of Total Cases

The total number of asymptomatic and symptomatic cases of COVID-19 were discerned in this study based on clinical testing programs – TATS for asymptomatic and CHS for symptomatic (see Section 2.4). Of the total 711 cases of COVID-19 that were reported via positive antigen and/or PCR clinical tests, the vast majority (79.2%) did not self-report and were therefore considered asymptomatic. We should note that this finding may be overestimated in the case that mildly symptomatic individuals did not self-report, which would categorize them as asymptomatic if tested and found positive via TATS. This high rate of asymptomatic cases may be due to the younger and perhaps healthier population surveyed in this study. However, the finding suggests that the number of people infected with SARs-CoV-2 in the U.S. may be vastly underestimated, which could have downstream effects for determining the immune population and herd immunity thresholds.

### 4.2 Viral Shedding Estimation

Fecal shedding rates of SARS-CoV-2 RNA were estimated from dorm wastewater samples using a modified equation from previous reports (Ahmed et al., 2020; Chavarria-Miró et al., 2021; Curtis et al., 2020). This calculation accounts for the number of infected people within a defined community, the amount of fecal material excreted, the shedding rate of infected individuals, the flow rate of the wastewater, and the viral load within the wastewater. Of these parameters, the fecal shedding rate of SARS-CoV-2 from infected individuals is the least known. Until this research, shedding rate estimations were based upon limited results that indicated a large variance across a small number of individuals (Wölfel et al., 2020). Shedding rates were further extrapolated from the 90^th^ percentile of this limited dataset and used to calculate the number of infected persons contributing to viral loads in wastewater (Ahmed et al., 2020; Chavarria-Miró et al., 2021; Curtis et al., 2020). However, models and predictions based on these percentile shedding rates are likely to be inaccurate without pairing wastewater results with clinical data.

In this current study, wastewater from student dorms with known defined communities was sampled and assayed for SARS-CoV-2 RNA. Wastewater samples that tested positive for viral RNA triggered targeted clinical testing of almost all residents within the specific dorm. Consequently, fecal shedding rates could be calculated since virus wastewater concentrations and the corresponding numbers of infected dorm residents were both known. These shedding rate estimations are reasonable since each dorm was a theoretically closed population, where nonresidents were prohibited from entering dorms, and individuals who were identified as positive were isolated within other facilities. This also prevented recounting of positive individuals and additional shedding on days subsequent to being counted as an infected individual, since persistent shedding occurs over multiple days (Cevik et al., 2020; Gupta et al., 2020). Therefore, the shedding rates calculated from the dorms represent incident infections only.

For estimating the number of individuals shedding and contributing to the viral load in wastewater samples, a 6-day range of clinical data was considered (See Section 2.5). Then, fecal shedding rates of SARS-CoV-2 RNA were extrapolated from wastewater viral loads in samples specific to UArizona dorms in which the exact number of infected persons was known. This estimation is based on a calculation that accounts for the number of infected persons within a defined community; the amount of fecal material excreted per person; the fecal shedding rate of viral RNA from infected individuals; the flow rate of the wastewater and the viral load within the wastewater (See Section 2.6).

The aggregation of wastewater and clinical data enabled quantification of SARS-CoV-2 fecal shedding rates per infected person. Mean shedding rates averaged over all dorms with positive wastewaters were 6.84 ± 0.77 log10 gc/g-feces based on the N1 gene and 7.74 ± 0.53 log10 gc/g-feces based on the N2 gene (Table 3). Statistical parameters included in Table 3 include number of samples (N), average values (Avg), standard deviation (Std dev), median (Med), minimum (Min), and maximum (Max). These statistics are important to establish the precision of the calculated shedding rates since many factors influence shedding rates. The calculated rates based on the incidence of both N1 and N2 are fairly consistent with reasonable calculated standard deviations. It is also important to note that mean rates are based on relatively large numbers of calculations: n = 60 (N1) and n = 32 (N2). Notably, N2 gene values were approximately 1-log greater than the N1 gene in feces.

While this paper develops population-level shedding rates, individual shedding rates may vary based on multiple factors such as sex, age, co-morbidities, socioeconomic status, and the severity of symptoms and duration of infection. The duration of shedding appears to be affected by the severity of the COVID-19 disease: asymptomatic (6 days) < mild symptoms (10 days) < moderate symptoms (12 days) < serious (14 days) < critical (32 days) (Chen et al., 2020). Recent reports also suggest that asymptomatic individuals had a shorter duration of viral shedding than pre-symptomatic individuals (Hue et al., 2020).

Our data assumes early infection shedding rates because those people would be identified within the 6-day period and removed from the dorm and brought into an isolated living quarter. However, it is possible that some individuals may have started and/or continued shedding viral RNA in feces outside of the assumed 6-day period. The shedding rate may also be slightly overestimated based on case capture definitions, as although the vast majority of individuals in each dorm were tested after positive wastewater sample results, a minority of residents were not tested. Shedding rates in this study are based on young university students who reside in dormitories and do not necessarily approximate the general population. Nevertheless, a recent study estimated the total number of active shedders from SARS-CoV-2 RNA levels in wastewater from a large metropolitan area similarly demonstrating that shedding resulted from a high proportion of asymptomatic individuals (Chavarria-Miró et al., 2021).

Some studies have reported similar initial shedding rates from symptomatic and asymptomatic infections (Lavezzo et al., 2020; Van Vinh Chau et al., 2020); whereas, other studies have found lower viral loads in asymptomatic cases (Han et al., 2020; Zhou et al., 2020). Also, some reports indicate that only approximately 50% of COVID-19 patients shed viral RNA in stool samples (Gupta et al., 2020; Medema et al., 2020a), suggesting the possibility that not all infected individuals shed viral RNA in feces. Regardless of these considerations, viral load in wastewater in this study were based on all individuals (symptomatic and asymptomatic) contributing to the sewage system. Therefore, fecal shedding estimates are general for defined communities of mixed clinical cases, while considering the high proportion of asymptomatic to symptomatic reported cases.

Lastly, it is important to note that no fecal samples were collected from individual COVID-19 patients directly. Therefore, results in this study are specific to wastewater from defined communities and is not intended to inform clinical data. Data and results from this study should be compared with clinical research and evaluations.

### 4.3 Concluding Remarks

This study may have significant implications for public health. The fecal shedding rate of SARS-CoV-2 RNA derived in this study can be utilized to estimate the total number of infections in a community based on wastewater viral concentrations. Knowledge of disease prevalence, especially as a leading indicator, can be used to assist communities in efficient resource allocation to prevent and contain COVID-19 outbreaks. This study also provides further understanding for the total number of cases that are symptomatic versus asymptomatic. Moving forward, the number of reported cases can provide context for estimating the number of cases that were asymptomatic and/or unreported. This, in turn, could have implications for understanding the proportion of individuals that has been exposed to COVID-19 and for understanding progress towards immunity within a community.

## Data Availability

Data is available upon request.

## Acknowledgements

The authors thank the UArizona Task Force, Amy Glicken, and Jeff Bliznick for their contributions. Financial support for the study was provided by the University of Arizona Campus Re-Entry Task Force. The IRB at the University of Arizona reviewed the study and verified that all data was de-identified and complied with the Human Subjects Protection.

