## Supplemental Material for "Enumerating asymptomatic COVID-19 cases and estimating SARS-CoV-2 fecal shedding rates via wastewater-based epidemiology"

Containing:
23 Pages
3 Tables

Table S1. Primer and probe nucleotide sequences for SARS-CoV-2 and HCoV 229E

| Primer or probe | Sequence (5’-3’) |
|---|---|
| 2019-nCoV_N1-F2019-nCoV_N1 | GACCCCAAAATCAGCGAAAT |
| 2019-nCoV_N1-R2019-nCoV_N1 | TCTGGTTACTGCCAGTTGAATCTG |
| 2019-nCoV_N1-P2019-nCoV_N1 | FAM-ACCCCGCATTACGTTTGGTGGACC-3IABkFQ* |
| 2019-nCoV_N2-F2019-nCoV_N2 | TTACAAACATTGGCCGCAAA |
| 2019-nCoV_N2-R2019-nCoV_N2 | GCGCGACATTCCGAAGAA |
| 2019-nCoV_N2-P2019-nCoV_N2 | FAM-ACAATTTGCCCCCAGCGCTTCAG-3IABkFQ* |
| 229E-FP | TTCCGACGTGCTCGAACTTT |
| 229E-RP | CCAACACGGTTGTGACAGTGA |
| 229E-TP | FAM-5-TCCTGAGGTCAATGCA-3-NFQ-MGB** |

* The FAM (6-carboxyfluorescein) quencher is BHQ-1 (Black Hole Quencher).

**The FAM quencher is a minor groove binder nonfluorescent quencher (MGBNFQ)**

Table S2. Performance characteristics of the RT-qPCR standard curves

| Assay | Estimated LoQ* | Cq | Slope | y-intercept | Dynamic range (copies/qPCR rxn) | Efficiency |
|---|---|---|---|---|---|---|
| N1 | 1.30E+02 | 35.25 | -3.437 | 45.52 | 20 - 2000000 | 0.95 |
| N2 | 1.01E+04 | 35.51 | -4.482 | 53.46 | 2000 - 2000000 | 0.65 |
| 229E | 3.86E+02 | 36.85 | -3.470 | 45.80 | 250 - 2500000 | 0.95 |

* LoQ = limit of quantification

Table S3: Wastewater and clinical data for SARS-CoV-2 at UArizona dormitories

A) Dorm A

| Sample Date | N1 Result[a] | N1 Concen-tration | N2 Result[a] | N2 Concen-tration | Occu-pancy | Clinical Tests[b] | Clinical Positives[b] | Sympto-matic[b] | Asympto-matic[b] | % Asymp-tomatic[b] |
|---|---|---|---|---|---|---|---|---|---|---|
| 8/17/2020 | Non-detect | - | Non-detect | - | 0 | 0 | 0 | 0 | 0 | |
| 8/19/2020 | Non-detect | - | Non-detect | - | 0 | 0 | 0 | 0 | 0 | |
| 8/24/2020 | Non-detect | - | Non-detect | - | 606 | 2 | 0 | 0 | 0 | |
| 8/26/2020 | Non-detect | - | Non-detect | - | 607 | 2 | 0 | 0 | 0 | |
| **8/31/2020** | **POSITIVE** | **3.73E+04** | **Non-detect** | **-** | **609** | **656** | **19** | **3** | **16** | **84.2%** |
| 9/1/2020 | Non-detect | - | Non-detect | - | 609 | 59 | 4 | 2 | 2 | 50.0% |
| **9/2/2020** | **POSITIVE** | **1.80E+04** | **Non-detect** | **-** | **608** | **646** | **19** | **3** | **16** | **84.2%** |
| 9/3/2020 | Non-detect | - | Non-detect | - | 608 | 380 | 6 | 1 | 5 | 83.3% |
| 9/4/2020 | Non-detect | - | Non-detect | - | 610 | 12 | 1 | 0 | 1 | 100.0% |
| **9/7/2020[c]** | **POSITIVE** | **7.42E+05** | **POSITIVE** | **8.42E+05** | **610** | **506** | **29** | **8** | **21** | **72.4%** |
| 9/9/2020 | Non-detect | - | Non-detect | - | 610 | 22 | 4 | 4 | 0 | 0.0% |
| **9/14/2020** | **POSITIVE** | **6.96E+04** | **POSITIVE** | **7.02E+05** | **610** | **494** | **83** | **19** | **64** | **77.1%** |
| **9/15/2020** | **POSITIVE** | **1.02E+05** | **POSITIVE** | **7.41E+05** | **610** | **494** | **83** | **19** | **64** | **77.1%** |
| **9/16/2020** | **POSITIVE** | **1.76E+05** | **Non-detect** | **-** | **610** | **429** | **57** | **7** | **50** | **87.7%** |
| 9/17/2020 | Non-detect | - | Non-detect | - | 610 | 66 | 5 | 2 | 3 | 60.0% |
| **9/18/2020** | **POSITIVE** | **3.41E+04** | **Non-detect** | **-** | **609** | **390** | **20** | **6** | **14** | **70.0%** |
| **9/21/2020** | **POSITIVE** | **7.22E+04** | **POSITIVE** | **7.93E+05** | **608** | **373** | **19** | **5** | **14** | **73.7%** |
| **9/23/2020** | **POSITIVE** | **4.20E+04** | **POSITIVE** | **8.23E+05** | **608** | **340** | **14** | **1** | **13** | **92.9%** |
| 9/28/2020 | Non-detect | - | Non-detect | - | 609 | 39 | 1 | 0 | 1 | 100.0% |
| **9/30/2020** | **POSITIVE** | **6.56E+04** | **Non-detect** | **-** | **609** | **166** | **1** | **0** | **1** | **100.0%** |
| **10/5/2020** | **POSITIVE** | **3.15E+04** | **Non-detect** | **-** | **605** | **158** | **2** | **0** | **2** | **100.0%** |
| 10/7/2020 | Non-detect | - | Non-detect | - | 606 | 38 | 0 | 0 | 0 | |
| **10/12/2020** | **POSITIVE** | **2.43E+03** | **Non-detect** | **-** | **607** | **165** | **4** | **0** | **4** | **100.0%** |
| 10/13/2020 | Non-detect | - | Non-detect | - | 606 | 22 | 0 | 0 | 0 | |
| **10/14/2020** | **POSITIVE** | **6.12E+03** | **Non-detect** | **-** | **605** | **137** | **2** | **0** | **2** | **100.0%** |
| 10/16/2020 | Non-detect | - | Non-detect | - | 606 | 33 | 0 | 0 | 0 | |

| | | | | | | | | | | |
|---|---|---|---|---|---|---|---|---|---|---|
| 10/19/2020 | Non-detect | - | Non-detect | - | 607 | 19 | 0 | 0 | 0 | |
| 10/21/2020 | Non-detect | - | Non-detect | - | 608 | 21 | 0 | 0 | 0 | |
| **10/28/2020** | **POSITIVE** | **1.39E+04** | **Non-detect** | **-** | **611** | **215** | **0** | **0** | **0** | |
| 11/2/2020 | Non-detect | - | Non-detect | - | 610 | 46 | 0 | 0 | 0 | |
| 11/4/2020 | Non-detect | - | Non-detect | - | 610 | 40 | 0 | 0 | 0 | |
| 11/9/2020 | Non-detect | - | Non-detect | - | 610 | 53 | 0 | 0 | 0 | |
| 11/11/2020 | Non-detect | - | Non-detect | - | 610 | 0 | 0 | 0 | 0 | |
| 11/16/2020 | Non-detect | - | Non-detect | - | 606 | 22 | 0 | 0 | 0 | |
| 11/18/2020 | Non-detect | - | Non-detect | - | 599 | 27 | 1 | 0 | 1 | 100.0% |
| 11/20/2020 | Non-detect | - | Non-detect | - | 590 | 25 | 0 | 0 | 0 | |
| | | | | | **Total** | **6097** | **374** | **80** | **294** | **78.6%** |

[a] 14 samples wastewater samples from Dorm A were positive for SARS-CoV-2 N1 and/or N2 genes as shown in blue. One positive sample (10/28) was omitted from viral shedding calculations due to 0 infected on those dates (Refer to Section 2.5).

[b] Clinical numbers for positive WW samples represent a 6-day range sum (Refer to Section 2.5). Clinical numbers for negative WW samples are day-of only. A negative wastewater sample did not trigger a response action to conduct clinical testing on residents; thus, there is limited clinical data for the days that wastewater was negative.

[c] Occupancy numbers for 9/6/20 and 9/7/20 are estimates based on the nearest date.

B) Dorm B

| Sample Date | N1 Result[a] | N1 Concen-tration | N2 Result[a] | N2 Concen-tration | Occu-pancy | Clinical Tests[b] | Clinical Positives[b] | Sympto-matic[b] | Asympto-matic[b] | % Asymp-tomatic[b] |
|---|---|---|---|---|---|---|---|---|---|---|
| 8/17/2020 | Non-detect | - | Non-detect | - | 0 | 0 | 0 | 0 | 0 | |
| 8/19/2020 | Non-detect | - | Non-detect | - | 0 | 0 | 0 | 0 | 0 | |
| 8/24/2020 | Non-detect | - | Non-detect | - | 341 | 0 | 0 | 0 | 0 | |
| 8/26/2020 | Non-detect | - | Non-detect | - | 340 | 0 | 0 | 0 | 0 | |
| 8/28/2020 | Non-detect | - | Non-detect | - | 342 | 4 | 0 | 0 | 0 | |
| **8/31/2020** | **POSITIVE** | **2.99E+04** | **POSITIVE** | **9.93E+05** | **341** | **358** | **15** | **2** | **13** | **86.7%** |
| 9/1/2020 | Non-detect | - | Non-detect | - | 342 | 308 | 11 | 1 | 10 | 90.9% |
| 9/2/2020 | Non-detect | - | Non-detect | - | 342 | 24 | 0 | 0 | 0 | |
| 9/7/2020[c] | Non-detect | - | Non-detect | - | 342 | 0 | 0 | 0 | 0 | |
| **9/9/2020** | **POSITIVE** | **2.01E+05** | **POSITIVE** | **6.99E+05** | **342** | **89** | **17** | **6** | **11** | **64.7%** |
| 9/10/2020 | Non-detect | - | Non-detect | - | 342 | 31 | 4 | 1 | 3 | 75.0% |
| 9/14/2020 | Non-detect | - | Non-detect | - | 342 | 239 | 41 | 2 | 39 | 95.1% |
| **9/16/2020** | **POSITIVE** | **2.44E+05** | **Non-detect** | **-** | **342** | **69** | **13** | **4** | **9** | **69.2%** |
| **9/21/2020** | **POSITIVE** | **5.61E+04** | **Non-detect** | **-** | **336** | **50** | **2** | **1** | **1** | **50.0%** |
| 9/28/2020 | Non-detect | - | Non-detect | - | 335 | 34 | 1 | 0 | 1 | 100.0% |
| **9/30/2020** | **POSITIVE** | **8.96E+03** | **Non-detect** | **-** | **335** | **78** | **1** | **0** | **1** | **100.0%** |
| **10/5/2020** | **POSITIVE** | **7.86E+03** | **Non-detect** | **-** | **332** | **66** | **0** | **0** | **0** | |
| 10/7/2020 | Non-detect | - | Non-detect | - | 332 | 20 | 0 | 0 | 0 | |
| 10/12/2020 | Non-detect | - | Non-detect | - | 332 | 14 | 0 | 0 | 0 | |
| 10/14/2020 | Non-detect | - | Non-detect | - | 331 | 13 | 0 | 0 | 0 | |
| 10/19/2020 | Non-detect | - | Non-detect | - | 330 | 21 | 0 | 0 | 0 | |
| 10/21/2020 | Non-detect | - | Non-detect | - | 330 | 13 | 1 | 1 | 0 | 0.0% |
| 10/21/2020 | Non-detect | - | Non-detect | - | 330 | 13 | 1 | 1 | 0 | 0.0% |
| 10/26/2020 | Non-detect | - | Non-detect | - | 329 | 14 | 0 | 0 | 0 | |
| **10/26/2020** | **POSITIVE** | **1.19E+04** | **Non-detect** | **-** | **329** | **124** | **0** | **0** | **0** | |
| 10/28/2020 | Non-detect | - | Non-detect | - | 329 | 27 | 0 | 0 | 0 | |
| 10/28/2020 | Non-detect | - | Non-detect | - | 329 | 27 | 0 | 0 | 0 | |

| | | | | | | | | | | |
|---|---|---|---|---|---|---|---|---|---|---|
| 11/2/2020 | Non-detect | - | Non-detect | - | 328 | 16 | 0 | 0 | 0 | |
| 11/2/2020 | Non-detect | - | Non-detect | - | 328 | 16 | 0 | 0 | 0 | |
| 11/4/2020 | Non-detect | - | Non-detect | - | 328 | 5 | 0 | 0 | 0 | |
| 11/4/2020 | Non-detect | - | Non-detect | - | 328 | 5 | 0 | 0 | 0 | |
| 11/9/2020 | Non-detect | - | Non-detect | - | 326 | 24 | 0 | 0 | 0 | |
| 11/9/2020 | Non-detect | - | Non-detect | - | 326 | 24 | 0 | 0 | 0 | |
| 11/16/2020 | Non-detect | - | Non-detect | - | 316 | 5 | 0 | 0 | 0 | |
| 11/16/2020 | Non-detect | - | Non-detect | - | 316 | 5 | 0 | 0 | 0 | |
| 11/18/2020 | Non-detect | - | Non-detect | - | 318 | 16 | 0 | 0 | 0 | |
| 11/18/2020 | Non-detect | - | Non-detect | - | 318 | 16 | 0 | 0 | 0 | |
| 11/20/2020 | Non-detect | - | Non-detect | - | 315 | 11 | 0 | 0 | 0 | |
| 11/20/2020 | Non-detect | - | Non-detect | - | 315 | 11 | 0 | 0 | 0 | |
| | | | | | **Total** | **1790** | **107** | **19** | **88** | **82.2%** |

[a] Seven wastewater samples from Dorm B were positive for SARS-CoV-2 N1 and/or N2 genes as shown in blue. Two positive samples (10/5 and 10/26) were omitted from viral shedding calculations due to 0 infected on those dates (Refer to Section 2.5).

[b] Clinical numbers for positive WW samples represent a 6-day range sum (Refer to Section 2.5). Clinical numbers for negative WW samples are day-of only. A negative wastewater sample did not trigger a response action to conduct clinical testing on residents; thus, there is limited clinical data for the days that wastewater was negative.

[c] Occupancy numbers for 9/6/20 and 9/7/20 are estimates based on the nearest date.

C) Dorm C

| Sample Date | N1 Result[a] | N1 Concen-tration | N2 Result[a] | N2 Concen-tration | Occu-pancy | Clinical Tests[b] | Clinical Positives[b] | Sympto-matic[b] | Asympto-matic[b] | % Asymp-tomatic[b] |
|---|---|---|---|---|---|---|---|---|---|---|
| 8/17/2020 | Non-detect | - | Non-detect | - | 0 | 0 | 0 | 0 | 0 | |
| 8/19/2020 | Non-detect | - | Non-detect | - | 0 | 0 | 0 | 0 | 0 | |
| 8/24/2020 | Non-detect | - | Non-detect | - | 123 | 1 | 0 | 0 | 0 | |
| 8/26/2020 | Non-detect | - | Non-detect | - | 123 | 1 | 0 | 0 | 0 | |
| 8/31/2020 | Non-detect | - | Non-detect | - | 123 | 0 | 0 | 0 | 0 | |
| 9/2/2020 | Non-detect | - | Non-detect | - | 123 | 6 | 3 | 0 | 3 | 100.0% |
| 9/4/2020 | Non-detect | - | Non-detect | - | 122 | 5 | 0 | 0 | 0 | |
| 9/7/2020[c] | Non-detect | - | Non-detect | - | 121 | 0 | 0 | 0 | 0 | |
| 9/9/2020 | Non-detect | - | Non-detect | - | 121 | 87 | 1 | 0 | 1 | 100.0% |
| 9/14/2020 | Non-detect | - | Non-detect | - | 121 | 4 | 1 | 0 | 1 | 100.0% |
| 9/16/2020 | Non-detect | - | Non-detect | - | 122 | 4 | 1 | 1 | 0 | 0.0% |
| 9/21/2020 | Non-detect | - | Non-detect | - | 122 | 4 | 1 | 1 | 0 | 0.0% |
| 9/28/2020 | Non-detect | - | Non-detect | - | 123 | 11 | 0 | 0 | 0 | |
| **9/30/2020** | **POSITIVE** | **8.96E+03** | **Non-detect** | **-** | **123** | **53** | **0** | **0** | **0** | |
| 10/5/2020 | Non-detect | - | Non-detect | - | 121 | 10 | 0 | 0 | 0 | |
| 10/7/2020 | Non-detect | - | Non-detect | - | 121 | 16 | 0 | 0 | 0 | |
| 10/12/2020 | Non-detect | - | Non-detect | - | 116 | 10 | 0 | 0 | 0 | |
| 10/14/2020 | Non-detect | - | Non-detect | - | 116 | 13 | 0 | 0 | 0 | |
| 10/19/2020 | Non-detect | - | Non-detect | - | 116 | 8 | 0 | 0 | 0 | |
| 10/21/2020 | Non-detect | - | Non-detect | - | 116 | 10 | 0 | 0 | 0 | |
| 10/26/2020 | Non-detect | - | Non-detect | - | 117 | 7 | 0 | 0 | 0 | |
| 10/28/2020 | Non-detect | - | Non-detect | - | 116 | 14 | 0 | 0 | 0 | |
| 11/2/2020 | Non-detect | - | Non-detect | - | 116 | 10 | 0 | 0 | 0 | |
| 11/4/2020 | Non-detect | - | Non-detect | - | 116 | 12 | 0 | 0 | 0 | |
| **11/9/2020** | **POSITIVE** | **4.13E+04** | **POSITIVE** | **2.04E+05** | **116** | **115** | **1** | **0** | **1** | **100.0%** |
| **11/11/2020** | **POSITIVE** | **1.94E+04** | **Non-detect** | **-** | **114** | **115** | **1** | **0** | **1** | **100.0%** |
| 11/16/2020 | Non-detect | - | Non-detect | - | 115 | 3 | 0 | 0 | 0 | |

| | | | | | | | | | | |
|---|---|---|---|---|---|---|---|---|---|---|
| 11/18/2020 | Non-detect | - | Non-detect | - | 115 | 5 | 0 | 0 | 0 | |
| 11/20/2020 | Non-detect | - | Non-detect | - | 114 | 3 | 0 | 0 | 0 | |
| | | | | | **Total** | **527** | **9** | **2** | **7** | **77.8%** |

[a] Three wastewater samples from Dorm C were positive for SARS-CoV-2 N1 and/or N2 genes as shown in blue. One positive sample (9/30) was omitted from viral shedding calculations due to 0 infected on those dates (Refer to Section 2.5).

[b] Clinical numbers for positive WW samples represent a 6-day range sum (Refer to Section 2.5). Clinical numbers for negative WW samples are day-of only. A negative wastewater sample did not trigger a response action to conduct clinical testing on residents; thus, there is limited clinical data for the days that wastewater was negative.

[c] Occupancy numbers for 9/6/20 and 9/7/20 are estimates based on the nearest date.

D) Dorm D

| Sample Date | N1 Result[a] | N1 Concen-tration | N2 Result[a] | N2 Concen-tration | Occu-pancy | Clinical Tests[b] | Clinical Positives[b] | Sympto-matic[b] | Asympto-matic[b] | % Asymp-tomatic[b] |
|---|---|---|---|---|---|---|---|---|---|---|
| 10/21/2020 | Non-detect | - | Non-detect | - | 582 | 19 | 0 | 0 | 0 | |
| 10/21/2020 | Non-detect | - | Non-detect | - | 582 | 19 | 0 | 0 | 0 | |
| 10/26/2020 | Non-detect | - | Non-detect | - | 579 | 11 | 0 | 0 | 0 | |
| 10/26/2020 | Non-detect | - | Non-detect | - | 579 | 11 | 0 | 0 | 0 | |
| 10/28/2020 | Non-detect | - | Non-detect | - | 579 | 42 | 1 | 0 | 1 | 100.0% |
| 10/28/2020 | Non-detect | - | Non-detect | - | 579 | 42 | 1 | 0 | 1 | 100.0% |
| 11/2/2020 | Non-detect | - | Non-detect | - | 569 | 25 | 0 | 0 | 0 | |
| 11/2/2020 | Non-detect | - | Non-detect | - | 569 | 25 | 0 | 0 | 0 | |
| 11/4/2020 | Non-detect | - | Non-detect | - | 566 | 17 | 0 | 0 | 0 | |
| 11/4/2020 | Non-detect | - | Non-detect | - | 566 | 17 | 0 | 0 | 0 | |
| 11/9/2020 | Non-detect | - | Non-detect | - | 557 | 39 | 0 | 0 | 0 | |
| 11/9/2020 | Non-detect | - | Non-detect | - | 557 | 39 | 0 | 0 | 0 | |
| 11/11/2020 | Non-detect | - | Non-detect | - | 555 | 0 | 0 | 0 | 0 | |
| 11/11/2020 | Non-detect | - | Non-detect | - | 555 | 0 | 0 | 0 | 0 | |
| 11/16/2020 | Non-detect | - | Non-detect | - | 544 | 11 | 0 | 0 | 0 | |
| **11/16/2020** | **Non-detect** | **-** | **POSITIVE** | **1.22E+05** | **544** | **128** | **1** | **0** | **1** | **100.0%** |
| 11/18/2020 | Non-detect | - | Non-detect | - | 540 | 18 | 0 | 0 | 0 | |
| 11/18/2020 | Non-detect | - | Non-detect | - | 540 | 18 | 0 | 0 | 0 | |
| 11/20/2020 | Non-detect | - | Non-detect | - | 530 | 18 | 0 | 0 | 0 | |
| 11/20/2020 | Non-detect | - | Non-detect | - | 530 | 18 | 0 | 0 | 0 | |
| | | | | | **Total** | **517** | **3** | **0** | **3** | **100.0%** |

[a] One wastewater sample from Dorm D was positive for SARS-CoV-2 N2 gene as shown in blue.

[b] Clinical numbers for positive WW samples represent a 6-day range sum (Refer to Section 2.5). Clinical numbers for negative WW samples are day-of only. A negative wastewater sample did not trigger a response action to conduct clinical testing on residents; thus, there is limited clinical data for the days that wastewater was negative.

[c] Occupancy numbers for 9/6/20 and 9/7/20 are estimates based on the nearest date.

E) Dorms E and F

| Sample Date | N1 Result[a] | N1 Concen-tration | N2 Result[a] | N2 Concen-tration | Occu-pancy | Clinical Tests[b] | Clinical Positives[b] | Sympto-matic[b] | Asympto-matic[b] | % Asymp-tomatic[b] |
|---|---|---|---|---|---|---|---|---|---|---|
| 8/18/2020 | Non-detect | - | Non-detect | - | 0 | 0 | 0 | 0 | 0 | |
| 8/20/2020 | Non-detect | - | Non-detect | - | 0 | 0 | 0 | 0 | 0 | |
| 8/25/2020 | Non-detect | - | Non-detect | - | 258 | 0 | 0 | 0 | 0 | |
| 8/27/2020 | Non-detect | - | Non-detect | - | 259 | 0 | 0 | 0 | 0 | |
| 9/1/2020 | Non-detect | - | Non-detect | - | 257 | 1 | 0 | 0 | 0 | |
| 9/3/2020 | Non-detect | - | Non-detect | - | 257 | 5 | 0 | 0 | 0 | |
| 9/8/2020 | Non-detect | - | Non-detect | - | 259 | 199 | 3 | 1 | 2 | 66.7% |
| 9/10/2020 | Non-detect | - | Non-detect | - | 260 | 12 | 0 | 0 | 0 | |
| **9/15/2020** | **POSITIVE** | **4.13E+04** | **Non-detect** | **-** | **260** | **53** | **9** | **5** | **4** | **44.4%** |
| 9/17/2020 | Non-detect | - | Non-detect | - | 260 | 4 | 2 | 2 | 0 | 0.0% |
| 9/18/2020 | Non-detect | - | Non-detect | - | 260 | 12 | 1 | 1 | 0 | 0.0% |
| **9/22/2020** | **POSITIVE** | **1.34E+04** | **Non-detect** | **-** | **260** | **94** | **2** | **2** | **0** | **0.0%** |
| 9/24/2020 | Non-detect | - | Non-detect | - | 260 | 10 | 1 | 1 | 0 | 0.0% |
| 9/29/2020 | Non-detect | - | Non-detect | - | 259 | 11 | 0 | 0 | 0 | |
| 10/1/2020 | Non-detect | - | Non-detect | - | 258 | 14 | 0 | 0 | 0 | |
| 10/6/2020 | Non-detect | - | Non-detect | - | 259 | 19 | 0 | 0 | 0 | |
| 10/8/2020 | Non-detect | - | Non-detect | - | 259 | 21 | 0 | 0 | 0 | |
| 10/13/2020 | Non-detect | - | Non-detect | - | 258 | 15 | 0 | 0 | 0 | |
| **10/15/2020** | **POSITIVE** | **7.74E+04** | **Non-detect** | **-** | **258** | **73** | **0** | **0** | **0** | |
| 10/16/2020 | Non-detect | - | Non-detect | - | 258 | 11 | 0 | 0 | 0 | |
| 10/20/2020 | Non-detect | - | Non-detect | - | 257 | 21 | 0 | 0 | 0 | |
| 10/22/2020 | Non-detect | - | Non-detect | - | 257 | 22 | 0 | 0 | 0 | |
| 10/27/2020 | Non-detect | - | Non-detect | - | 256 | 40 | 1 | 0 | 1 | 100.0% |
| 10/29/2020 | Non-detect | - | Non-detect | - | 256 | 26 | 1 | 0 | 1 | 100.0% |
| 11/3/2020 | Non-detect | - | Non-detect | - | 255 | 18 | 0 | 0 | 0 | |
| 11/5/2020 | Non-detect | - | Non-detect | - | 254 | 8 | 0 | 0 | 0 | |
| 11/10/2020 | Non-detect | - | Non-detect | - | 249 | 18 | 0 | 0 | 0 | |

| | | | | | | | | | | |
|---|---|---|---|---|---|---|---|---|---|---|
| 11/12/2020 | Non-detect | - | Non-detect | - | 248 | 19 | 0 | 0 | 0 | |
| 11/17/2020 | Non-detect | - | Non-detect | - | 240 | 24 | 0 | 0 | 0 | |
| 11/19/2020 | Non-detect | - | Non-detect | - | 240 | 15 | 0 | 0 | 0 | |
| | | | | | **Total** | **765** | **20** | **12** | **8** | **40.0%** |

[a] Three wastewater samples from combined Dorms E and F were positive for SARS-CoV-2 N1 gene as shown in blue. One positive sample (10/15) was omitted from viral shedding calculations due to 0 infected on those dates (Refer to Section 2.5).

[b] Clinical numbers for positive WW samples represent a 6-day range sum (Refer to Section 2.5). Clinical numbers for negative WW samples are day-of only. A negative wastewater sample did not trigger a response action to conduct clinical testing on residents; thus, there is limited clinical data for the days that wastewater was negative.

[c] Occupancy numbers for 9/6/20 and 9/7/20 are estimates based on the nearest date.

F) Dorm G

| Sample Date | N1 Result[a] | N1 Concen-tration | N2 Result[a] | N2 Concen-tration | Occu-pancy | Clinical Tests[b] | Clinical Positives[b] | Sympto-matic[b] | Asympto-matic[b] | % Asymp-tomatic[b] |
|---|---|---|---|---|---|---|---|---|---|---|
| 10/21/2020 | Non-detect | - | Non-detect | - | 222 | 18 | 0 | 0 | 0 | |
| 10/26/2020 | Non-detect | - | Non-detect | - | 223 | 17 | 0 | 0 | 0 | |
| 10/28/2020 | Non-detect | - | Non-detect | - | 223 | 13 | 1 | 0 | 1 | 100.0% |
| 11/2/2020 | Non-detect | - | Non-detect | - | 221 | 27 | 0 | 0 | 0 | |
| 11/4/2020 | Non-detect | - | Non-detect | - | 221 | 20 | 0 | 0 | 0 | |
| 11/9/2020 | Non-detect | - | Non-detect | - | 221 | 12 | 0 | 0 | 0 | |
| 11/11/2020 | Non-detect | - | Non-detect | - | 221 | 0 | 0 | 0 | 0 | |
| 11/16/2020 | Non-detect | - | Non-detect | - | 222 | 6 | 0 | 0 | 0 | |
| 11/18/2020 | Non-detect | - | Non-detect | - | 222 | 4 | 0 | 0 | 0 | |
| 11/20/2020 | Non-detect | - | Non-detect | - | 222 | 4 | 0 | 0 | 0 | |
| | | | | | **Total** | **121** | **1** | **0** | **1** | **100.0%** |

[a] No wastewater samples from Dorm G were positive for SARS-CoV-2 N1 and/or N2 genes.

[b] Clinical numbers for positive WW samples represent a 6-day range sum (Refer to Section 2.5). Clinical numbers for negative WW samples are day-of only. A negative wastewater sample did not trigger a response action to conduct clinical testing on residents; thus, there is limited clinical data for the days that wastewater was negative.

[c] Occupancy numbers for 9/6/20 and 9/7/20 are estimates based on the nearest date.

G) Dorms H and I

| Sample Date | N1 Result[a] | N1 Concen-tration | N2 Result[a] | N2 Concen-tration | Occu-pancy | Clinical Tests[b] | Clinical Positives[b] | Sympto-matic[b] | Asympto-matic[b] | % Asymp-tomatic[b] |
|---|---|---|---|---|---|---|---|---|---|---|
| 8/18/2020 | Non-detect | - | Non-detect | - | 0 | 0 | 0 | 0 | 0 | |
| 8/20/2020 | Non-detect | - | Non-detect | - | 0 | 0 | 0 | 0 | 0 | |
| 8/25/2020 | Non-detect | - | Non-detect | - | 373 | 2 | 0 | 0 | 0 | |
| 8/27/2020 | Non-detect | - | Non-detect | - | 374 | 2 | 0 | 0 | 0 | |
| **9/1/2020** | **POSITIVE** | **2.22E+05** | **Non-detect** | **-** | **375** | **225** | **9** | **4** | **5** | **55.6%** |
| **9/2/2020** | **POSITIVE** | **3.44E+04** | **Non-detect** | **-** | **373** | **215** | **9** | **4** | **5** | **55.6%** |
| 9/3/2020 | Non-detect | - | Non-detect | - | 373 | 18 | 0 | 0 | 0 | |
| **9/4/2020** | **POSITIVE** | **6.45E+05** | **POSITIVE** | **4.11E+06** | **373** | **334** | **9** | **2** | **7** | **77.8%** |
| **9/6/2020[c]** | **POSITIVE** | **7.27E+03** | **Non-detect** | **-** | **374** | **197** | **13** | **3** | **10** | **76.9%** |
| **9/8/2020** | **POSITIVE** | **3.42E+04** | **Non-detect** | **-** | **374** | **330** | **21** | **4** | **17** | **81.0%** |
| 9/9/2020 | Non-detect | - | Non-detect | - | 374 | 9 | 1 | 0 | 1 | 100.0% |
| **9/10/2020** | **POSITIVE** | **6.38E+04** | **Non-detect** | **-** | **374** | **250** | **30** | **6** | **24** | **80.0%** |
| **9/11/2020** | **POSITIVE** | **8.70E+04** | **Non-detect** | **-** | **374** | **276** | **35** | **8** | **27** | **77.1%** |
| **9/14/2020** | **POSITIVE** | **2.00E+05** | **POSITIVE** | **2.76E+06** | **372** | **274** | **33** | **8** | **25** | **75.8%** |
| **9/15/2020** | **POSITIVE** | **2.46E+04** | **Non-detect** | **-** | **372** | **274** | **33** | **8** | **25** | **75.8%** |
| 9/17/2020 | Non-detect | - | Non-detect | - | 372 | 30 | 5 | 3 | 2 | 40.0% |
| **9/22/2020** | **POSITIVE** | **6.19E+04** | **Non-detect** | **-** | **371** | **91** | **7** | **1** | **6** | **85.7%** |
| **9/24/2020** | **POSITIVE** | **3.33E+05** | **Non-detect** | **-** | **371** | **155** | **6** | **1** | **5** | **83.3%** |
| 9/25/2020 | Non-detect | - | Non-detect | - | 371 | 18 | 4 | 1 | 3 | 75.0% |
| 9/29/2020 | Non-detect | - | Non-detect | - | 372 | 31 | 0 | 0 | 0 | |
| 10/1/2020 | Non-detect | - | Non-detect | - | 371 | 26 | 0 | 0 | 0 | |
| 10/6/2020 | Non-detect | - | Non-detect | - | 371 | 31 | 0 | 0 | 0 | |
| **10/8/2020** | **POSITIVE** | **2.33E+04** | **Non-detect** | **-** | **370** | **133** | **1** | **0** | **1** | **100.0%** |
| 10/9/2020 | Non-detect | - | Non-detect | - | 371 | 17 | 0 | 0 | 0 | |
| 10/13/2020 | Non-detect | - | Non-detect | - | 372 | 35 | 0 | 0 | 0 | |
| 10/15/2020 | Non-detect | - | Non-detect | - | 371 | 30 | 0 | 0 | 0 | |
| 10/20/2020 | Non-detect | - | Non-detect | - | 366 | 25 | 0 | 0 | 0 | |

| | | | | | | | | | | |
|---|---|---|---|---|---|---|---|---|---|---|
| 10/22/2020 | Non-detect | - | Non-detect | - | 365 | 24 | 0 | 0 | 0 | |
| 10/27/2020 | Non-detect | - | Non-detect | - | 364 | 50 | 0 | 0 | 0 | |
| 10/29/2020 | Non-detect | - | Non-detect | - | 363 | 31 | 0 | 0 | 0 | |
| 11/3/2020 | Non-detect | - | Non-detect | - | 363 | 31 | 0 | 0 | 0 | |
| 11/5/2020 | Non-detect | - | Non-detect | - | 363 | 33 | 0 | 0 | 0 | |
| 11/10/2020 | Non-detect | - | Non-detect | - | 360 | 29 | 0 | 0 | 0 | |
| 11/12/2020 | Non-detect | - | Non-detect | - | 360 | 27 | 0 | 0 | 0 | |
| **11/17/2020** | **POSITIVE** | **2.26E+05** | **Non-detect** | **-** | **345** | **145** | **0** | **0** | **0** | |
| 11/19/2020 | Non-detect | - | Non-detect | - | 340 | 43 | 0 | 0 | 0 | |
| | | | | | **Total** | **3441** | **216** | **53** | **163** | **75.5%** |

[a] 13 wastewater samples from combined Dorms H and I were positive for SARS-CoV-2 N1 and/or N2 genes as shown in blue. One positive sample (10/8) was omitted from viral shedding calculations due to 0 infected on those dates (Refer to Section 2.5).

[b] Clinical numbers for positive WW samples represent a 6-day range sum (Refer to Section 2.5). Clinical numbers for negative WW samples are day-of only. A negative wastewater sample did not trigger a response action to conduct clinical testing on residents; thus, there is limited clinical data for the days that wastewater was negative.

[c] Occupancy numbers for 9/6/20 and 9/7/20 are estimates based on the nearest date.

H) Dorm J

| Sample Date | N1 Result[a] | N1 Concen-tration | N2 Result[a] | N2 Concen-tration | Occu-pancy | Clinical Tests[b] | Clinical Positives[b] | Sympto-matic[b] | Asympto-matic[b] | % Asymp-tomatic[b] |
|---|---|---|---|---|---|---|---|---|---|---|
| 8/18/2020 | Non-detect | - | Non-detect | - | 0 | 0 | 0 | 0 | 0 | |
| 8/20/2020 | Non-detect | - | Non-detect | - | 0 | 0 | 0 | 0 | 0 | |
| 8/25/2020 | Non-detect | - | Non-detect | - | 424 | 1 | 0 | 0 | 0 | |
| 8/27/2020 | Non-detect | - | Non-detect | - | 423 | 2 | 0 | 0 | 0 | |
| **9/1/2020** | **POSITIVE** | **5.90E+04** | **Non-detect** | **-** | **420** | **409** | **14** | **1** | **13** | **92.9%** |
| **9/2/2020** | **Non-detect** | **-** | **POSITIVE** | **5.95E+05** | **419** | **404** | **14** | **1** | **13** | **92.9%** |
| 9/3/2020 | Non-detect | - | Non-detect | - | 419 | 19 | 0 | 0 | 0 | |
| 9/4/2020 | Non-detect | - | Non-detect | - | 417 | 360 | 11 | 1 | 10 | 90.9% |
| **9/8/2020** | **POSITIVE** | **2.68E+04** | **Non-detect** | **-** | **419** | **347** | **23** | **4** | **19** | **82.6%** |
| **9/9/2020** | **POSITIVE** | **8.19E+04** | **POSITIVE** | **6.99E+05** | **419** | **347** | **23** | **4** | **19** | **82.6%** |
| **9/10/2020** | **POSITIVE** | **6.88E+05** | **POSITIVE** | **7.80E+05** | **419** | **451** | **39** | **5** | **34** | **87.2%** |
| **9/11/2020** | **POSITIVE** | **1.14E+06** | **POSITIVE** | **9.33E+05** | **418** | **459** | **45** | **8** | **37** | **82.2%** |
| **9/14/2020** | **POSITIVE** | **6.23E+04** | **POSITIVE** | **7.02E+05** | **417** | **435** | **42** | **9** | **33** | **78.6%** |
| **9/15/2020** | **POSITIVE** | **1.28E+05** | **POSITIVE** | **7.41E+05** | **417** | **435** | **42** | **9** | **33** | **78.6%** |
| **9/17/2020** | **POSITIVE** | **1.40E+08** | **Non-detect** | **-** | **417** | **331** | **24** | **3** | **21** | **87.5%** |
| **9/18/2020** | **POSITIVE** | **4.55E+04** | **POSITIVE** | **7.41E+05** | **416** | **319** | **19** | **1** | **18** | **94.7%** |
| **9/22/2020** | **POSITIVE** | **1.09E+05** | **POSITIVE** | **7.27E+05** | **415** | **304** | **14** | **0** | **14** | **100.0%** |
| **9/24/2020** | **POSITIVE** | **2.57E+05** | **POSITIVE** | **1.02E+06** | **415** | **286** | **11** | **1** | **10** | **90.9%** |
| **9/25/2020** | **POSITIVE** | **4.44E+05** | **Non-detect** | **-** | **413** | **309** | **11** | **1** | **10** | **90.9%** |
| **9/29/2020** | **Non-detect** | **-** | **POSITIVE** | **5.53E+05** | **412** | **309** | **14** | **2** | **12** | **85.7%** |
| **10/1/2020** | **POSITIVE** | **2.73E+04** | **Non-detect** | **-** | **412** | **232** | **10** | **1** | **9** | **90.0%** |
| 10/6/2020 | Non-detect | - | Non-detect | - | 403 | 15 | 0 | 0 | 0 | |
| 10/8/2020 | Non-detect | - | Non-detect | - | 402 | 36 | 1 | 0 | 1 | 100.0% |
| 10/9/2020 | Non-detect | - | Non-detect | - | 403 | 7 | 0 | 0 | 0 | |
| 10/13/2020 | Non-detect | - | Non-detect | - | 394 | 20 | 0 | 0 | 0 | |
| 10/15/2020 | Non-detect | - | Non-detect | - | 393 | 31 | 0 | 0 | 0 | |
| 10/20/2020 | Non-detect | - | Non-detect | - | 389 | 15 | 0 | 0 | 0 | |

| | | | | | | | | | | |
|---|---|---|---|---|---|---|---|---|---|---|
| 10/22/2020 | Non-detect | - | Non-detect | - | 388 | 15 | 0 | 0 | 0 | |
| 10/27/2020 | Non-detect | - | Non-detect | - | 388 | 44 | 0 | 0 | 0 | |
| 10/29/2020 | Non-detect | - | Non-detect | - | 387 | 34 | 0 | 0 | 0 | |
| **11/3/2020** | **POSITIVE** | **2.03E+04** | **POSITIVE** | **2.14E+05** | **384** | **209** | **0** | **0** | **0** | |
| 11/5/2020 | Non-detect | - | Non-detect | - | 383 | 105 | 0 | 0 | 0 | |
| **11/10/2020** | **POSITIVE** | **1.21E+04** | **Non-detect** | **-** | **377** | **120** | **1** | **0** | **1** | **100.0%** |
| 11/12/2020 | Non-detect | - | Non-detect | - | 372 | 20 | 0 | 0 | 0 | |
| **11/17/2020** | **POSITIVE** | **3.59E+05** | **POSITIVE** | **1.16E+05** | **364** | **85** | **0** | **0** | **0** | |
| 11/19/2020 | Non-detect | - | Non-detect | - | 361 | 15 | 0 | 0 | 0 | |
| | | | | | **Total** | **6530** | **358** | **51** | **307** | **85.8%** |

[a] 18 wastewater samples from Dorm J were positive for SARS-CoV-2 N1 and/or N2 genes as shown in blue. Two positive samples (11/3 and 11/17) were omitted from viral shedding calculations due to 0 infected on those dates (Refer to Section 2.5).

[b] Clinical numbers for positive WW samples represent a 6-day range sum (Refer to Section 2.5). Clinical numbers for negative WW samples are day-of only. A negative wastewater sample did not trigger a response action to conduct clinical testing on residents; thus, there is limited clinical data for the days that wastewater was negative.

[c] Occupancy numbers for 9/6/20 and 9/7/20 are estimates based on the nearest date.

I) Dorm K

| Sample Date | N1 Result[a] | N1 Concen-tration | N2 Result[a] | N2 Concen-tration | Occu-pancy | Clinical Tests[b] | Clinical Positives[b] | Sympto-matic[b] | Asympto-matic[b] | % Asymp-tomatic[b] |
|---|---|---|---|---|---|---|---|---|---|---|
| 8/18/2020 | Non-detect | - | Non-detect | - | 0 | 0 | 0 | 0 | 0 | |
| 8/20/2020 | Non-detect | - | Non-detect | - | 0 | 0 | 0 | 0 | 0 | |
| **8/25/2020** | **Non-detect** | **-** | **POSITIVE** | **1.61E+05** | **317** | **592** | **3** | **1** | **2** | **66.7%** |
| **8/26/2020** | **POSITIVE** | **3.84E+05** | **POSITIVE** | **1.06E+06** | **317** | **591** | **3** | **1** | **2** | **66.7%** |
| **8/26/2020** | **POSITIVE** | **3.74E+05** | **POSITIVE** | **1.06E+06** | **317** | **591** | **3** | **1** | **2** | **66.7%** |
| **8/26/2020** | **Non-detect** | **-** | **POSITIVE** | **1.06E+06** | **317** | **591** | **3** | **1** | **2** | **66.7%** |
| **8/26/2020** | **POSITIVE** | **1.73E+05** | **POSITIVE** | **1.06E+06** | **317** | **591** | **3** | **1** | **2** | **66.7%** |
| **8/26/2020** | **POSITIVE** | **3.77E+05** | **POSITIVE** | **1.06E+06** | **317** | **591** | **3** | **1** | **2** | **66.7%** |
| 8/27/2020 | Non-detect | - | Non-detect | - | 317 | 26 | 0 | 0 | 0 | |
| 8/27/2020 | Non-detect | - | Non-detect | - | 317 | 26 | 0 | 0 | 0 | |
| 8/28/2020 | Non-detect | - | Non-detect | - | 319 | 88 | 0 | 0 | 0 | |
| **8/29/2020** | **POSITIVE** | **1.00E+04** | **POSITIVE** | **9.93E+05** | **319** | **315** | **3** | **2** | **1** | **33.3%** |
| 8/30/2020 | Non-detect | - | Non-detect | - | 319 | 0 | 0 | 0 | 0 | |
| **9/1/2020** | **POSITIVE** | **8.68E+03** | **POSITIVE** | **7.39E+05** | **321** | **45** | **3** | **3** | **0** | **0.0%** |
| 9/2/2020 | Non-detect | - | Non-detect | - | 324 | 6 | 2 | 2 | 0 | 0.0% |
| **9/3/2020** | **POSITIVE** | **1.40E+04** | **Non-detect** | **-** | **324** | **23** | **3** | **3** | **0** | **0.0%** |
| 9/4/2020 | Non-detect | - | Non-detect | - | 325 | 12 | 0 | 0 | 0 | |
| 9/6/2020[c] | Non-detect | - | Non-detect | - | 325 | 0 | 0 | 0 | 0 | |
| 9/8/2020 | Non-detect | - | Non-detect | - | 325 | 198 | 8 | 0 | 8 | 100.0% |
| **9/10/2020** | **POSITIVE** | **3.24E+04** | **Non-detect** | **-** | **325** | **74** | **15** | **4** | **11** | **73.3%** |
| 9/11/2020 | Non-detect | - | Non-detect | - | 326 | 18 | 0 | 0 | 0 | |
| **9/15/2020** | **Non-detect** | **-** | **POSITIVE** | **7.41E+05** | **328** | **305** | **27** | **6** | **21** | **77.8%** |
| **9/17/2020** | **POSITIVE** | **6.74E+04** | **Non-detect** | **-** | **328** | **123** | **13** | **5** | **8** | **61.5%** |
| **9/18/2020** | **POSITIVE** | **1.01E+04** | **Non-detect** | **-** | **328** | **136** | **14** | **5** | **9** | **64.3%** |
| **9/22/2020** | **Non-detect** | **-** | **POSITIVE** | **7.27E+05** | **325** | **291** | **20** | **5** | **15** | **75.0%** |
| 9/24/2020 | Non-detect | - | Non-detect | - | 325 | 84 | 4 | 0 | 4 | 100.0% |
| **9/29/2020** | **Non-detect** | **-** | **POSITIVE** | **5.53E+05** | **325** | **179** | **3** | **0** | **3** | **100.0%** |

| | | | | | | | | | | |
|---|---|---|---|---|---|---|---|---|---|---|
| **10/1/2020** | **POSITIVE** | **1.17E+04** | **Non-detect** | **-** | **325** | **130** | **2** | **0** | **2** | **100.0%** |
| 10/6/2020 | Non-detect | - | Non-detect | - | 325 | 42 | 0 | 0 | 0 | |
| 10/8/2020 | Non-detect | - | Non-detect | - | 325 | 26 | 0 | 0 | 0 | |
| **10/9/2020** | **POSITIVE** | **4.74E+04** | **Non-detect** | **-** | **325** | **120** | **1** | **0** | **1** | **100.0%** |
| 10/12/2020 | Non-detect | - | Non-detect | - | 327 | 29 | 1 | 0 | 1 | 100.0% |
| 10/13/2020 | Non-detect | - | Non-detect | - | 327 | 39 | 0 | 0 | 0 | |
| 10/15/2020 | Non-detect | - | Non-detect | - | 326 | 42 | 0 | 0 | 0 | |
| **10/20/2020** | **POSITIVE** | **4.95E+04** | **Non-detect** | **-** | **326** | **259** | **0** | **0** | **0** | |
| **10/22/2020** | **POSITIVE** | **1.32E+05** | **Non-detect** | **-** | **326** | **259** | **0** | **0** | **0** | |
| 10/27/2020 | Non-detect | - | Non-detect | - | 325 | 43 | 0 | 0 | 0 | |
| 10/29/2020 | Non-detect | - | Non-detect | - | 325 | 51 | 0 | 0 | 0 | |
| **11/3/2020** | **POSITIVE** | **2.07E+04** | **POSITIVE** | **3.12E+05** | **327** | **272** | **0** | **0** | **0** | |
| 11/5/2020 | Non-detect | - | Non-detect | - | 326 | 119 | 0 | 0 | 0 | |
| 11/10/2020 | Non-detect | - | Non-detect | - | 325 | 48 | 0 | 0 | 0 | |
| 11/12/2020 | Non-detect | - | Non-detect | - | 322 | 55 | 0 | 0 | 0 | |
| 11/17/2020 | Non-detect | - | Non-detect | - | 324 | 48 | 0 | 0 | 0 | |
| 11/19/2020 | Non-detect | - | Non-detect | - | 321 | 50 | 0 | 0 | 0 | |
| 8/31/2020 | Non-detect | - | Non-detect | - | 319 | 5 | 0 | 0 | 0 | |
| | | | | | **Total** | **7133** | **137** | **41** | **96** | **70.1%** |

[a] 20 wastewater samples from Dorm K were positive for SARS-CoV-2 N1 and/or N2 genes as shown in blue. Three positive samples (10/20, 10/22, and 11/3) were omitted from viral shedding calculations due to 0 infected on those dates (Refer to Section 2.5).

[b] Clinical numbers for positive WW samples represent a 6-day range sum (Refer to Section 2.5). Clinical numbers for negative WW samples are day-of only. A negative wastewater sample did not trigger a response action to conduct clinical testing on residents; thus, there is limited clinical data for the days that wastewater was negative.

[c] Occupancy numbers for 9/6/20 and 9/7/20 are estimates based on the nearest date.

J) Dorm L

| Sample Date | N1 Result[a] | N1 Concen-tration | N2 Result[a] | N2 Concen-tration | Occu-pancy | Clinical Tests[b] | Clinical Positives[b] | Sympto-matic[b] | Asympto-matic[b] | % Asymp-tomatic[b] |
|---|---|---|---|---|---|---|---|---|---|---|
| 8/17/2020 | Non-detect | - | Non-detect | - | 0 | 0 | 0 | 0 | 0 | |
| 8/19/2020 | Non-detect | - | Non-detect | - | 0 | 0 | 0 | 0 | 0 | |
| 8/24/2020 | Non-detect | - | Non-detect | - | 74 | 0 | 0 | 0 | 0 | |
| 8/26/2020 | Non-detect | - | Non-detect | - | 74 | 0 | 0 | 0 | 0 | |
| 8/31/2020 | Non-detect | - | Non-detect | - | 75 | 1 | 0 | 0 | 0 | |
| 9/2/2020 | Non-detect | - | Non-detect | - | 75 | 0 | 0 | 0 | 0 | |
| 9/7/2020[c] | Non-detect | - | Non-detect | - | 75 | 0 | 0 | 0 | 0 | |
| 9/9/2020 | Non-detect | - | Non-detect | - | 75 | 57 | 0 | 0 | 0 | |
| 9/14/2020 | Non-detect | - | Non-detect | - | 74 | 0 | 0 | 0 | 0 | |
| 9/16/2020 | Non-detect | - | Non-detect | - | 74 | 9 | 0 | 0 | 0 | |
| 9/21/2020 | Non-detect | - | Non-detect | - | 74 | 4 | 0 | 0 | 0 | |
| 9/23/2020 | Non-detect | - | Non-detect | - | 74 | 5 | 0 | 0 | 0 | |
| 9/28/2020 | Non-detect | - | Non-detect | - | 76 | 8 | 0 | 0 | 0 | |
| 9/30/2020 | Non-detect | - | Non-detect | - | 76 | 12 | 0 | 0 | 0 | |
| 10/5/2020 | Non-detect | - | Non-detect | - | 76 | 6 | 0 | 0 | 0 | |
| 10/7/2020 | Non-detect | - | Non-detect | - | 76 | 9 | 0 | 0 | 0 | |
| 10/12/2020 | Non-detect | - | Non-detect | - | 75 | 5 | 0 | 0 | 0 | |
| 10/14/2020 | Non-detect | - | Non-detect | - | 75 | 7 | 0 | 0 | 0 | |
| 10/19/2020 | Non-detect | - | Non-detect | - | 74 | 7 | 0 | 0 | 0 | |
| 10/21/2020 | Non-detect | - | Non-detect | - | 74 | 11 | 0 | 0 | 0 | |
| 10/26/2020 | Non-detect | - | Non-detect | - | 73 | 6 | 0 | 0 | 0 | |
| 10/28/2020 | Non-detect | - | Non-detect | - | 73 | 9 | 0 | 0 | 0 | |
| 11/2/2020 | Non-detect | - | Non-detect | - | 74 | 5 | 0 | 0 | 0 | |
| 11/4/2020 | Non-detect | - | Non-detect | - | 74 | 7 | 0 | 0 | 0 | |
| 11/9/2020 | Non-detect | - | Non-detect | - | 73 | 8 | 0 | 0 | 0 | |
| 11/11/2020 | Non-detect | - | Non-detect | - | 73 | 0 | 0 | 0 | 0 | |
| 11/16/2020 | Non-detect | - | Non-detect | - | 73 | 3 | 0 | 0 | 0 | |

| | | | | | | | | | | |
|---|---|---|---|---|---|---|---|---|---|---|
| 11/18/2020 | Non-detect | - | Non-detect | - | 73 | 7 | 0 | 0 | 0 | |
| 11/20/2020 | Non-detect | - | Non-detect | - | 73 | 3 | 0 | 0 | 0 | |
| | | | | | **Total** | **189** | **0** | **0** | **0** | **-** |

[a] No wastewater samples from Dorm L were positive for SARS-CoV-2 N1 and/or N2 genes as shown in blue.

[b] Clinical numbers for positive WW samples represent a 6-day range sum (Refer to Section 2.5). Clinical numbers for negative WW samples are day-of only. A negative wastewater sample did not trigger a response action to conduct clinical testing on residents; thus, there is limited clinical data for the days that wastewater was negative.

[c] Occupancy numbers for 9/6/20 and 9/7/20 are estimates based on the nearest date.

K) Dorm M

| Sample Date | N1 Result[a] | N1 Concen-tration | N2 Result[a] | N2 Concen-tration | Occu-pancy | Clinical Tests[b] | Clinical Positives[b] | Sympto-matic[b] | Asympto-matic[b] | % Asymp-tomatic[b] |
|---|---|---|---|---|---|---|---|---|---|---|
| 10/22/2020 | Non-detect | - | Non-detect | - | 129 | 8 | 0 | 0 | 0 | |
| 10/27/2020 | Non-detect | - | Non-detect | - | 129 | 12 | 0 | 0 | 0 | |
| 10/29/2020 | Non-detect | - | Non-detect | - | 129 | 8 | 0 | 0 | 0 | |
| 11/3/2020 | Non-detect | - | Non-detect | - | 129 | 11 | 0 | 0 | 0 | |
| 11/5/2020 | Non-detect | - | Non-detect | - | 129 | 18 | 0 | 0 | 0 | |
| **11/10/2020** | **POSITIVE** | **6.61E+04** | **Non-detect** | **-** | **128** | **44** | **0** | **0** | **0** | |
| 11/12/2020 | Non-detect | - | Non-detect | - | 128 | 4 | 0 | 0 | 0 | |
| 11/17/2020 | Non-detect | - | Non-detect | - | 128 | 23 | 0 | 0 | 0 | |
| **11/19/2020** | **POSITIVE** | **6.32E+05** | **POSITIVE** | **2.19E+06** | **128** | **40** | **0** | **0** | **0** | |
| | | | | | **Total** | **168** | **0** | **0** | **0** | **-** |

[a] Two wastewater samples from Dorm M were positive for SARS-CoV-2 N1 and/or N2 genes as shown in blue. Both samples (11/10 and 11/19) were omitted from viral shedding calculations due to 0 infected on those dates (Refer to Section 2.5).

[b] Clinical numbers for positive WW samples represent a 6-day range sum (Refer to Section 2.5). Clinical numbers for negative WW samples are day-of only. A negative wastewater sample did not trigger a response action to conduct clinical testing on residents; thus, there is limited clinical data for the days that wastewater was negative.

[c] Occupancy numbers for 9/6/20 and 9/7/20 are estimates based on the nearest date.
